# Cross-Program Secondary Analyses and Public Health Innovation: The RADx Data Hub as a Resource for Integrated COVID-19 Research

**DOI:** 10.1101/2025.11.26.25341110

**Authors:** Peter W. Rose, Thais Rivas, Nader Mehri, Oswaldo Alonso Lozoya, Liesl Jeffers-Francis, Keriayn N. Smith, Marquea D. King, Mete U. Akdogan, Jasmine Snipe, Brandy Farlow, Michael A. Keller, Mark A. Musen, Stephanie Suber, Ashok Krishnamurthy

## Abstract

The COVID-19 pandemic highlighted the need for timely, standardized research data to address future health challenges. From the pandemic, there has been a plethora of diverse data, including patient demographics, clinical outcomes, biospecimen information, and technology development frameworks. Yet, much of these data have been fragmented and without clear provenance and standardization. To overcome these limitations, the National Institutes of Health (NIH) established the RADx Data Hub (Data Hub) as a centralized platform to promote Rapid Acceleration of Diagnostics (RADx) data distribution under the Findable, Accessible, Interoperable, and Reusable (FAIR) data principles. The Data Hub aggregated diverse COVID-19 diagnostic and clinical datasets into a harmonized repository that supports data integration, quality control, and secondary research. We independently tested Data Hub biomedical data management and data analysis capabilities to investigate its utility in viral monitoring, health disparity research, biomarker identification, and precarity modeling. We found that the Data Hub’s accessibility enabled efficient secondary research, including diagnostic and clinical data reuse. This supports repository use in accelerating infectious disease research and studying disease outcomes. Further, the Data Hub’s centralized, FAIR-aligned structure exemplifies a sustainable data stewardship model, extending beyond COVID-19 to addressing future public health emergencies, while strengthening the capacity to do reproducible science within broader biomedical research.

## Introduction

The COVID-19 pandemic exposed critical research gaps that precluded the efficient data collection, integration, and analysis needed to enable a rapid public health response. To assist, the National Institutes of Health (NIH) launched the Rapid Acceleration of Diagnostics (RADx) Initiative (1). This initiative, comprised of four initial programs: RADx Radical (RADx-rad), RADx Underserved Populations (RADx-UP), RADx Tech, and RADx Digital Health Technologies (RADx DHT), diverse yet complementary programs spanning experimental diagnostics, health disparities research, technology development, and digital health solutions (1,2). Specifically, RADx-rad focused on novel diagnostic approaches for improved COVID-19 detection, including innovative assays and targets, while RADx-UP focused on understanding and addressing COVID-19 health disparities (1). RADx Tech accelerated COVID-19 test development, validation, and commercialization, and RAD DHT generated and integrated digital health data from diverse sources (1,2).

The final program, the RADx Data Hub (Data Hub; radxdatahub.nih.gov), was formed to centralize and standardize RADx data from more than 180 studies and 1700 data files (2), aggregating vast and valuable COVID-19 data to enable secondary research and accelerate diagnostic innovations and public health impacts (2). Within the Data Hub, structured approaches were developed to validate, deidentify, and integrate data, supporting findable, accessible, interoperable, and reusable (FAIR) data principles (2). For users, the Data Hub provided harmonized data access within a secure environment equipped with appropriate analytical tools, including statistical software and Jupyter notebooks, to facilitate secondary intra- and cross-study RADx program data investigations (2).

The goal behind this organization and access was to eliminate common primary data collection time and resource constraints faced by researchers, to facilitate enriched exploration of research questions and foster efficient hypothesis testing to accelerate COVID-19 (and related) knowledge generation. The Data Hub’s harmonization framework categorized data elements into tiers reflecting their degree of standardization and shared use across multiple studies (2). This structure integrates diverse datasets and enables comparative analysis (including Tier 1, universal data elements across all studies) to support comprehensive foundational information. Additional tiered variables represent common, program-specific data elements, tailored to certain study subsets or non-harmonized, study-specific variables (2). For example, metadata on RADx-rad specimen types, analytes, detection approaches, and biological targets were systematically catalogued across diagnostic sub-projects (2,3).

The goal of these Data Hub functionalities is to support broad secondary research to advance understanding of COVID-19 and similar diseases. This includes population-level viral monitoring through diagnostic and environmental surveillance data, tracking viral/variant emergence and spread to inform public health responses, and insights into vulnerable community testing access and infection outcome inequities. Further, biomarker identification is an important use case for diagnostics and prognostics, with integrable datasets for researchers to identify biological signatures applicable for disease detection and monitoring. Predictive modeling with harmonized Hub datasets should also support statistical and machine learning approaches to forecast outbreak dynamics, providing insights into disease trajectories that can strengthen preparedness and response planning.

Here, we independently tested Data Hub capacity to address empirical research questions, seeking to understand the utility of this and similar data repositories to enable secondary research and data validation, both for COVID-19 and other infectious and non-infectious diseases.

## Materials and Methods

### Broad utility testing

RADx-rad data were accessed through the dbGaP portal (https://dbgap.ncbi.nlm.nih.gov/home). We identified 44 RADx-rad studies with available datasets in the Data Hub (https://radxdatahub.nih.gov/) and imported them into a Data Hub workspace configured with AWS SageMaker Studio. The workspace, provisioned with 100 GB of disk space and 16 GB of memory, provided direct data access. Each study appeared in a folder labeled by its dbGaP accession number, containing data and dictionary files (CSV) and metadata (JSON). The metadata included key identifiers such as subproject names, which were used for aggregation and cross-study analysis. Analyses were performed in Jupyter Notebooks within a reproducible CONDA environment. The notebooks, environment specification, and RADx-rad data dictionaries annotated with tier information (https://github.com/radxrad/common-data-elements) are publicly available on GitHub (https://github.com/radxrad/radx-analyzer).

### Wastewater analyses

For comparative wastewater studies, sampling frame and locations were: i) NC A&T (Greensboro, NC): grab or time-composite wastewater samples collected from dormitory sewer laterals and central campus interceptors on scheduled sampling days (2021–2023), with sampling twice per week during spring and fall semesters and once per week during summer; and ii) Miami University (Oxford, OH): wastewater samples from community-scale sewersheds with sampling once per week, available via the RADx Infectious Disease (RADx-rad) program and accessed through the Data Hub. Samples from NC A&T were transported on ice, concentrated, extracted, and quantified for SARS-CoV-2 RNA copies/L by RT-qPCR using standard assays (e.g., N1/N2), with appropriate calibration and controls (4). For NC A&T data aggregation, per-date mean across dorms was computed (zeros treated as missing if flagged as below limit of detection (LoD)). For Miami University data (source Data Hub) aggregation, per-date mean across sites was computed from processed RADx data. All time series are shown on a log10 scale to accommodate multi-order-of-magnitude variation.

Examined variant windows included dominant variant periods (Alpha, Delta, Omicron BA.1, BA.5, XBB), annotated as light-grey bands using widely accepted date ranges for U.S. circulation (5,6). For site comparability, NC A&T dorm-level catchments represented smaller, higher-resolution populations, whereas Miami University sewersheds represented broader community populations, affecting signal amplitude and variance.

### Health Disparity Analyses

We accessed six Data Hub datasets focused on community interventions to increase COVID-19 testing and vaccination among Latino populations in California, Maryland, New York, and Florida. Search filters included the terms “Latino” and “Latinx,” “community intervention,” “COVID-19,” and “social determinants of health.” Participants completed structured questionnaires assessing demographic, socioeconomic, and health-related characteristics, including education level, employment status, sex, age group, and health insurance coverage. COVID-19 positivity was determined through self-reporting or verified diagnostic test results. Additional items captured perceptions and concerns about the COVID-19 vaccine, including safety, development speed, effectiveness, and trust in government or healthcare systems. The measures used included: i) COVID-19 Positivity Rate: Defined as the proportion of participants who reported a positive COVID-19 test result within each subgroup (education, employment, insurance type, sex, and age); ii) Vaccine Concerns: Categorized as safety/side effects, rapid development, doubts about effectiveness, distrust of government, distrust of healthcare, or religious/moral concerns; and iii) Sociodemographic Variables: Education (less than high school, high school, some college, bachelor’s, graduate), employment status (full-time, part-time, unemployed, homemaker, self-employed, student), and insurance type (uninsured, public, private). For data management, all six source files were combined, and variable names were aligned across datasets to enable cross-study comparison. Data cleaning procedures addressed inconsistent entries, including “missing,” “other,” and “prefer not to answer” responses. Qualitative data were extracted from interviews and focus groups within each study, capturing detailed information on community outreach efforts, vaccine concerns, and testing and vaccination barriers. Both quantitative and qualitative measures were used to evaluate uptake, participation rates, and participant perceptions of vaccination and testing programs. Data were last accessed on November 5, 2025. Authors did not have access to data that could identify individual study participants.

Qualitative data were analyzed thematically to identify recurring barriers, facilitators, and perceptions related to COVID-19 testing and vaccination. Quantitative measures of participation and vaccination uptake were summarized descriptively to contextualize cross-site intervention effectiveness. Mixed-methods data integration provided a comprehensive understanding of how social determinants of health, such as education, employment, and insurance access, affected COVID-19 outcomes within Latino communities.

### Retrospective characterization of RNA-seq library complexity

Raw sequencing volumes: Based on reported information retrieved from the published study, specimen RNA-seq data exhibited a broad range of total sequencing volumes, between 1.5M-160M reads sequenced per sample (median: 32.4M, IQR: 24.7M-39.4M) and transcriptome read coverage levels between 11.4x-46.1x (median: 29.8x, IQR: 26.1x-33.5x). These values corresponded to rates of sequenced reads per 1x transcriptome coverage equivalent between 117K-3.72M reads across specimens (median: 1.05M, IQR: 843K-1.27M).

Sequencing saturation metrics: Further inspection of the gene expression matrix supplied to the Data Hub revealed substantial changes in the amount and dispersion for the total counts of reads aligned uniquely to transcripts, between 28.6K-37.9M counts per sample (median: 4.30M, IQR: 2.10M-6.85M) which corresponds to 1.50-1.28K fold difference in volume-to-aligned read rates (median: 5.88, IQR: 4.50-11.2). Normalized volume-to-aligned read rates represent a wide range of sequencing saturation levels across specimens in the expression matrix from the original study. Interestingly, these rates corresponded to estimates for effective aligned depth, i.e., the relation between total numbers of aligned reads and the 1x transcriptome coverage equivalent rate in a specimen, between 0.01x-15.8x (median: 4.78x, IQR: 2.42x-7.05x) implying that the overall gene expression matrix obtained from the original study, after unique sequenced reads alignment and deduplication, is also sparse and under-sampled in terms of number of genes detected and gene read counts per specimen, respectively.

### Parametric focusing for specimen selection from unbalanced and sparse RNA-seq data

To handle the original Data Hub study’s sparse expression matrix (7), we selected a reduced search space of parametrically segregated, variably expressed (facultative) genes, i.e., gene knee plot. This approach shows improved performance under unsupervised clustering analysis compared to approaches that include all genes regardless of their total aligned read tallies (8). The SALSA analytical workflow is a statistically agnostic method for biomarker extraction from single-cell RNA-seq data; it segregates single cell representations (barcodes) agnostically, based on latent patterns of shared gene expression and is tailored to process unary expression data from large combinations of individual low-complexity libraries sequenced beyond saturation. Further, to mitigate disparate library complexity saturation in gene expression analysis impacts, we adopted parametric sweeping of rank-ordered total read counts per barcode, i.e., specimen knee plot, a technique from scRNA-seq data processing by which barcodes from putative cell multiplets (with an “excess” volume of reads), cell singlets (with a “sufficient” volume of reads), and fractional cell debris (with a “shallow” volume of reads) are segregated by their total read counts under a probabilistic mixture parametric model (8).

Transcriptome readout efficiency aggregate metrics supported our parametric-based segregation of rank-ordered specimens based on total aligned reads, identifying subsets of sequenced specimens in which gene expression information, which flows from total reads sequenced to net aligned reads and transcriptome equivalent coverage, may lose correspondence. The pool of 142 sufficient specimens (2.6M-19M total aligned reads per sample) kept for unsupervised classification and differential expression analysis showed matching ranges for transcriptome readout efficiency metrics that tracked commensurately along the bioinformatics workflow: volume-to-aligned fold differences between 1.50-19.5 (median: 5.01, IQR: 4.34-6.66) and effective aligned depth rates between 1.59x-15.8x (median: 6.37x, IQR: 4.54x-7.72x). In contrast, 6 excess specimens were discarded from further analysis (21M-38M total aligned reads per sample), displaying narrow transcriptome readout efficiency metric ranges, suggesting overwhelming representation of RNA from highly expressed genes: volume-to-aligned fold differences between 3.60-4.47 (median: 3.99, IQR: 3.72-4.45) and effective aligned depth rates between 8.32x-11.1x (median: 9.58x, IQR: 8.78x-10.46x). 69 shallow specimens (29K-2.5M total aligned reads per sample) were also discarded from further analysis, as they presented wide and mismatched transcriptome readout efficiency metric ranges by orders of magnitude, characteristic of data from oversaturated and clonal sequencing of low-complexity libraries: volume-to-aligned fold differences between 3.74-1.28K (median: 16.1, IQR: 8.61-27.5) and effective aligned depth rates between 0.01x-5.95x (median: 1.65x, IQR: 0.83x-2.46x).

### Precarity modeling

RADx-UP promoted equitable COVID-19 testing, vaccination, and care access among disproportionately affected communities; it was the source of Precarity modeling data. The Data Hub’s Study Explorer and Metadata Specification tools assisted in selecting nine RADx-UP projects, applying filters for study population, design, and variable availability to identify datasets containing standardized Common Data Elements (CDEs) relevant to household hardship and vaccination. Together, these projects contributed de-identified data from 5,397 adults across diverse settings including rural, urban, and Tribal communities, representing historically underrepresented populations in biomedical research.

Six household challenge variables were selected (including limited access to healthcare, unstable housing, food insecurity, lack of clean water, difficulty obtaining medications, and limited transportation) and modeled as observed indicators in a CFA to derive a latent Precarity construct. Weighted least squares mean and variance-adjusted estimation (WLSMV) was used for ordinal data, and model fit was assessed using χ², CFI, TLI, and RMSEA indices. Factor scores from the validated model were then used in a multivariable logistic regression predicting COVID-19 vaccination uptake, adjusting for education, age, sex, and insurance coverage. Analyses were conducted in R (v4.3) using the lavaan and glm packages.

## Results

### 1) Utility of RADx Study Data

#### 1.1. Description of RADx-rad Studies used in Utility Testing

To test general Data Hub data utility, we analyzed 44 RADx-rad studies accessed through the Data Hub and grouped into three research areas: i) wastewater-based surveillance, ii) Multisystem Inflammatory Syndrome in Children (MIS-C) diagnosis, and iii) novel diagnostic methods development. Collectively, these studies span community-level SARS-CoV-2 surveillance, clinical diagnostics, and point-of-care technology innovation.

Six subprojects focused on wastewater monitoring for population-level SARS-CoV-2 (and its variants) detection. These studies evaluated viral RNA detection in environmental samples, examined temporal trends, and benchmarked wastewater signals against case incidence data, demonstrating wastewater surveillance as a scalable public health monitoring tool. Eight Predicting Viral-Associated Inflammatory Disease Severity in Children with Laboratory Diagnostics and Artificial Intelligence (PreVAIL-kIds) subprojects addressed MIS-C diagnostic and biomarker discovery. These studies investigated biomarkers, clinical characteristics, and outcomes prediction, providing insights into pediatric complications of SARS-CoV-2 infection in vulnerable populations. The remaining 30 subprojects developed innovative diagnostic technologies across six thematic areas (Table 1).

**Table 1.** Diagnostic Technologies.

| Technology | Description |
| --- | --- |
| SCENT (Electronic Noses) | VOC-based breath analysis for SARS-CoV-2 detection |
| Chemosensory Testing | At-home smell tests for rapid, low-cost screening |
| Exosome-Based Technologies | Microfluidic saliva assays for viral proteins and RNA, antibodies, and host exosome signatures |
| Novel Biosensing | Home-based diagnostics and breathalyzers for sensitive, rapid testing |
| Automatic Detection and Tracing | Electrochemical and mechanical sensors, touch-based detection, aerosolized virus monitoring, and privacy-preserving contact tracing |
| Multimodal Surveillance | Integrated systems such as smart masks, dialysis facility monitoring, and multimodal data streams for early detection and population surveillance |

#### 1.2. Analysis of Data Elements Across Studies

Each dataset included a data dictionary with standardized study variables and defined names, types, and definitions, ensuring consistent representation and interpretation across studies. The 46 Tier 1 data elements, defined by NIH as describing patient-level metadata (demographics, disability status, health history, mental health status, and COVID-related symptoms), were harmonized across all RADx projects to enable cross-study analyses. Tier 2 elements, focused on technology development, are harmonized across RADx-rad subprojects, while Tier 3 data elements comprise project-specific variables that could not be further harmonized.

Both the relative proportions among Tiers 1-3 and the number of data elements within each tier varied considerably across studies (Fig 1A-B). Although 46 Tier 1 data elements were defined, not all were available or applicable to every study (Fig 1B). Tier 2 data elements showed broader coverage across diagnostic technology studies, whereas Tier 3 data elements reflected project-specific measurements. The PreVAIL-kIds (MIS-C) studies, in particular, exhibited substantial diversity, using primarily Tier 3 data elements. This variation highlights the breadth of RADx-rad data within the Data Hub and opportunities for further harmonization to enhance cross-study comparability.

**Figure 1.**
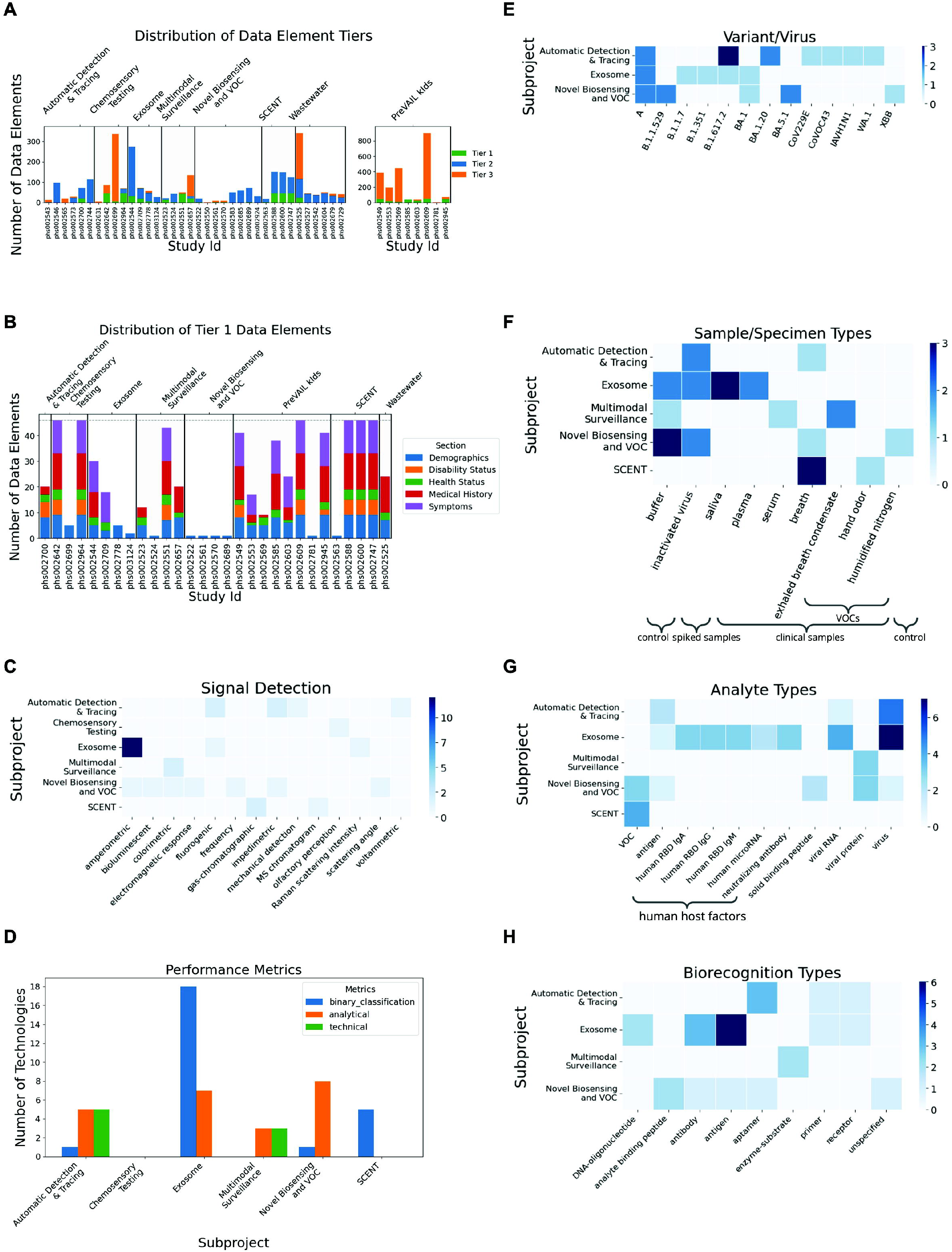
Inventory of study characteristics across NIH RADx-rad projects. Overview of data element tiers, specimen types, analytes, detection methods, and biological targets reported across RADx-rad subprojects. A. Distribution of Tier 1-3 data elements across studies, grouped by subproject. Tier 1 elements represent patient-level metadata harmonized across RADx projects; Tier 2 elements represent technology-development variables harmonized across RADx-rad subprojects; Tier 3 elements are project-specific and not further harmonized. B. Breakdown of Tier 1 data elements by category, including demographics, disability status, health history, mental health, and symptoms. Although 46 harmonized Tier 1 elements were defined, not all were reported in every study. C. Types and frequencies of signal-detection methods employed across diagnostic subprojects. D. Metrics reported to evaluate diagnostic performance, including sensitivity, specificity, predictive values, percent agreement, and area under the ROC curve (AUC), as well as analytical and technical measures such as limit of detection (LoD) and turnaround time. E. SARS-CoV-2 variants tested across subprojects, including Alpha (B.1.1.7), Beta (B.1.351), Delta (B.1.617.2), Omicron (BA lineages), and other variants of concern, as well as common respiratory viruses (e.g., human coronaviruses and influenza) included for cross-reactivity testing. F. Specimen types analyzed across subprojects, including saliva, breath, blood/serum, and nasal swabs. G. Classes of analytes detected by RADx-rad diagnostic methods, including viral proteins, RNA, antibodies, and host biomarkers. H. Biorecognition mechanisms used across subprojects, such as antibodies, aptamers, enzyme-based reporters, and volatile-organic-compound (VOC) recognition systems.

#### 1.3. Analysis of Diagnostic Method Development Studies

RADx-rad diagnostic projects encompassed wide-ranging technologies, including chromatographic, electrochemical, fluorescent, mechanical, spectroscopic, and olfactory modalities (Fig 1C). This diversity reflects complementary signal generation strategies, designed to optimize sensitivity and robustness across diagnostic contexts. Projects reported multiple performance metrics categories (Fig 1D), including Binary classification metrics: sensitivity, specificity, predictive values, percent agreement, and area under the ROC curve (AUC); Analytical metrics: limit of LoD, limit of quantitation (LoQ), and limit of blank (LoB), used to benchmark assay sensitivity; and technical metrics: cost per test and turnaround time, addressing practical field deployment considerations.

Despite variable reporting, several platforms achieved >90% sensitivity and specificity, with low LoDs enabling early infection detection. Cross-reactivity testing against influenza and seasonal coronaviruses confirmed diagnostic specificity, while validation across Alpha, Beta, Delta, Omicron, and other variants demonstrated robustness in dynamic epidemiological contexts (Fig 1E).

Diagnostic studies employed diverse specimen types, including saliva, breath, blood/serum, and nasal swabs (Fig 1F). Saliva and breath were most common because they are non-invasive and suitable for at-home testing. Analytes included viral spike proteins, proteases, and antibodies, enabling detection across multiple stages of infection (Fig 1G). Biorecognition approaches spanned antibodies, aptamers, enzyme-based reporters, and VOC recognition systems (Fig 1H).

### 2. Wastewater SARS-CoV-2 dynamics

#### 2.1. Selection of Data Hub wastewater study and external comparator

To assess whether Data Hub-housed wastewater data may contextualize campus-level surveillance, we paired (i) an external dataset from North Carolina Agricultural and Technical State University (NC A&T) with (ii) a community-scale wastewater dataset from Miami University, available in the Data Hub (“University of Miami SARS-CoV-2 wastewater surveillance” study). The Miami University dataset was clearly documented, sampled repeatedly over the COVID-19 period, and represented a broad, community-scale sewershed, suitable as a reference for the higher-resolution NC A&T dorm/interceptor data. This pairing enabled testing of cross-site comparability and the added value of Data Hub resources for campuses already collecting local wastewater data.

#### 2.2. Semester-linked wastewater dynamics across campuses

After harmonizing both datasets to daily means (NC A&T: per-date mean across dorms, zeros treated as missing if below LoD; Miami: per-date mean across sites from the processed RADx table), both campuses showed semester-linked trends with surges near term starts (Fig 2A), consistent with prior findings that campus population shifts appear in wastewater signals (4).

**Figure 2.**
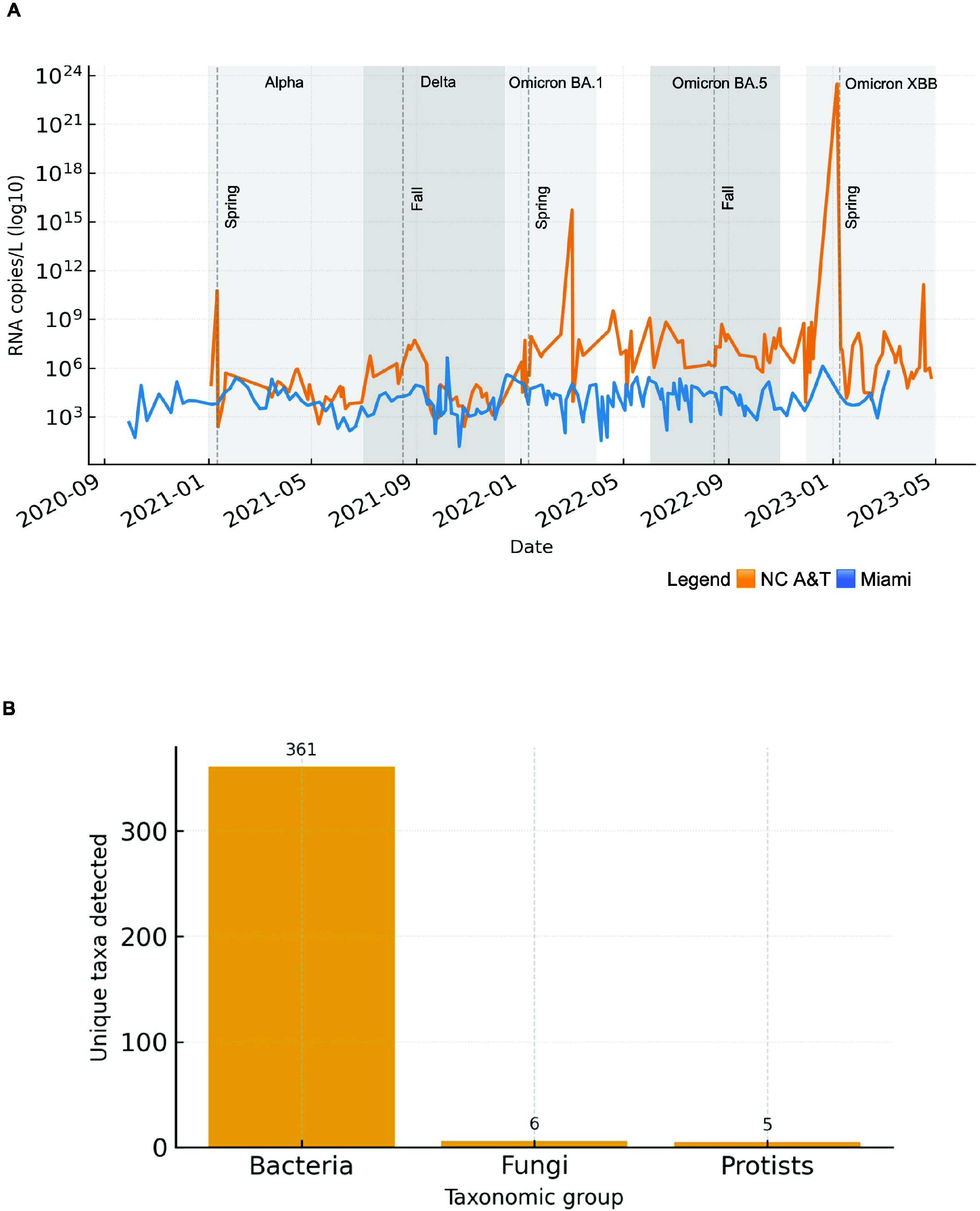
Wastewater SARS-CoV-2 dynamics with campus microbiome context. A. Miami vs NC A&T wastewater RNA over time (2021–2023). NC A&T = orange, Miami = blue; y-axis = RNA copies/L (log10). Dashed vertical lines mark semester starts. Gray bands indicate dominant variant waves (Alpha, Delta, Omicron BA.1, BA.5, XBB). NC A&T = mean across dorms per date; Miami = mean per date across sites. B. NC A&T wastewater microbial diversity (metagenomics). Unique taxa detected: Bacteria 361, Fungi 6, Protists 5.

Signal amplitude and sharpness differed by catchment scale (defined as the population and sewer infrastructure feeding a given sampling point). Miami University (community-scale mean) exhibited higher-amplitude, sharper peaks around campus returns, while NC A&T (dorm-level mean) showed a lower baseline with episodic increases (4). These differences are expected, because larger, community-scale sewersheds integrate more people and introductions, while dormitory sewer laterals represent smaller, higher-resolution catchments with more stochastic variation.

#### 2.3. Alignment with SARS-CoV-2 variant periods

To interpret these peaks, we overlaid dominant U.S. variant periods (Alpha, Delta, Omicron BA.1, BA.5, XBB) as contextual, light-grey bands (Fig 2A). The largest SARS-CoV-2 RNA concentration increases (i.e., the highest peaks in the wastewater signal) at both campuses fell within Delta and Omicron windows, matching clinical reports that Omicron lineages spread faster and showed immune escape even with shorter clinical lead times (5,6). This concordance indicates that Data Hub wastewater data can serve as a temporal reference for campus data (Fig 2A).

#### 2.4. Local vs. regional resolution from paired datasets

A key objective was to determine whether the paired datasets could distinguish local from regional signals. NC A&T’s dorm-level signals provided localized, actionable signals tied to specific campus areas, supporting targeted communication or testing. In contrast, the Data Hub series for Miami University provided broad, regional context and external validation, showing whether campus increases reflected broader community rise. Together, these findings decompose the wastewater signal into campus trends relative to the surrounding community (4).

#### 2.5. Biosurveillance capacity from metagenomics

Finally, to determine whether the workflow could support endpoints beyond SARS-CoV-2, we summarized NC A&T metagenomics results. Campus wastewater contained 361 bacterial, 6 fungal, and 5 protist taxa (Fig 2B), demonstrating that the same sampling and data-integration workflow can support broader biosurveillance and environmental health monitoring (9). This finding complements the comparative wastewater analysis and confirms that campus wastewater also captures diverse microbial signatures, enabling future expansion to RSV, influenza, and anti-microbial resistance (AMR) monitoring.

### 3. Analysis of COVID-19 in Diverse Populations

To assess the capacity to address COVID-19-related health disparities and evaluate community-based intervention impacts on COVID-19 testing and vaccine uptake in Latino populations, we used data from six NIH RADx studies across California, Maryland, New York, and Florida. We combined quantitative and qualitative data, standardized variables, and assessed structural and behavioral factors to evaluate engagement barriers and facilitators.

#### 3.1. COVID-19 Positivity by Sex and Education Level in Latino Participants

COVID-19 positivity varied by both sex and education across Latino participants in the six RADx studies (Fig 3A). Participants with less than a high school education had the highest positivity rates (60% of women and 57% of men), indicating increased vulnerability among individuals with limited access to stable employment, healthcare, and preventive resources (Fig 3A).

**Figure 3.**
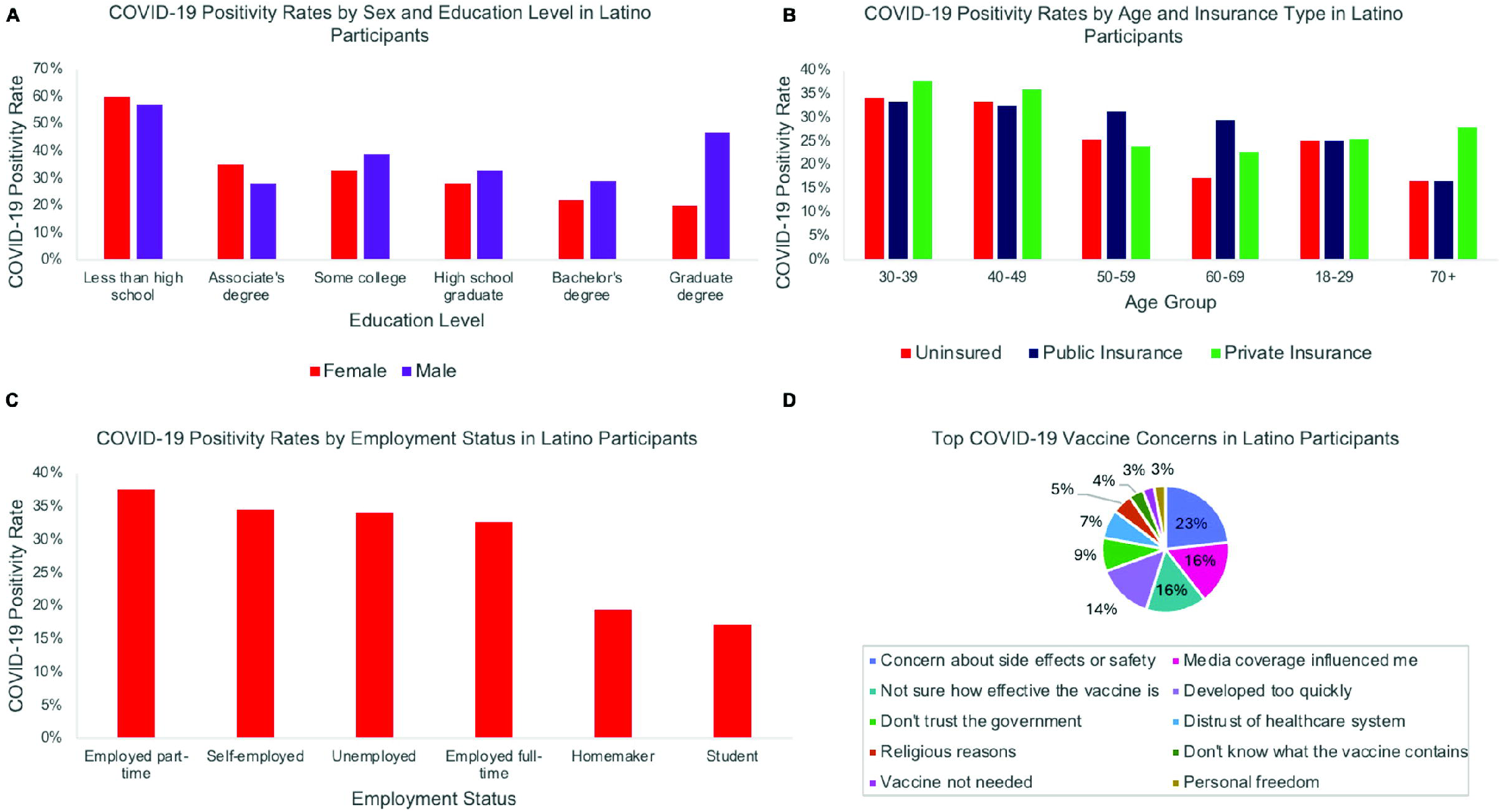
COVID-19 testing and vaccine concerns among Latino participants. A. COVID-19 positivity rates stratified by sex (male, female) and education level (less than high school, high school graduate, some college or higher). B. COVID-19 positivity rates stratified by age group (18-29, 30-49, 50+) and insurance type (insured, uninsured). C. COVID-19 positivity rates stratified by employment status (full-time, part-time, unemployed, essential worker). D. The most commonly reported COVID-19 vaccine concerns, including safety, efficacy, development speed, and institutional trust. Data were pooled from six RADx studies of Latino communities across California, Maryland, New York, and Florida. Percentages reflect the proportion of participants within each subgroup reporting positive COVID-19 tests or specific vaccine concerns.

Positivity decreased with higher educational attainment. Among participants with a bachelor’s degree, 22% of women and 29% of men tested positive; among those with graduate degrees, rates fell to 20% for women and rose to 47% for men. The unexpectedly higher rate among graduate-educated men likely reflects a smaller sample size (low *n*) and potential occupational heterogeneity.

Participants with associate’s degrees or some college education showed moderate positivity rates (28-39%), possibly influenced by job type and workforce participation. Men with “some college” exhibited slightly higher rates (39%) than women (33%), possibly due to employment in construction, manufacturing, transportation, and service roles where physical distancing and remote work were limited. Women consistently showed lower positivity rates across most educational levels, possibly relating to differences in occupational exposure.

#### 3.2. COVID-19 Positivity by Age and Insurance Type

Participants aged 30-39 years had the highest positivity rates across insurance types, reaching 37.8% among those with private insurance, 34.1% among the uninsured, and 33.3% among those with public insurance (Fig 3B). This pattern suggests that early- and mid-career adults faced substantial exposure regardless of insurance coverage. Among participants aged 40-49, positivity remained elevated across insurance groups (32.5-36.0%), then declined with age. In the 50-59 group, positivity fell to 23.8-31.2%, with the highest among those on public insurance, potentially reflecting underlying health vulnerabilities and increased testing access through public programs.

Participants aged 60-69 showed a pronounced disparity: 29.5% positivity among those with public insurance compared to 17.3% for the uninsured and 22.7% with private insurance, suggesting differential testing access and chronic disease burden among older Medicare or Medicaid beneficiaries. In the 70+ group, positivity was lowest among publicly insured participants (16.7%) but increased to 27.8% for those with private insurance. Among young adults (18–29), positivity was consistent across all insurance types (∼25%), suggesting that social behaviors and occupational exposure, rather than coverage, drove infection risks (Fig 3B).

Overall, insurance coverage did not uniformly protect against infection; exposure risks, occupational context, and healthcare access jointly influenced outcomes. Age-stratified disparities highlight the need for targeted outreach and prevention strategies, especially for middle-aged Latino adults balancing high exposure risk and variable healthcare access. These findings reflect the intersection of economic precarity and inadequate healthcare access, where uninsured, working-age adults experienced delayed testing and reduced preventative care.

#### 3.3. COVID-19 Positivity by Employment Status in Latino Participants

Participants who were employed part-time (37.6%) and self-employed (34.5%) had the highest positivity rates (Fig 3C), possibly due to inconsistent workplace protections, lack of paid sick leave, and dependence on multiple income sources that increase daily contact. Unemployed participants (34.1%) showed comparably high rates, likely influenced by prior exposure in high-contact jobs or ongoing socioeconomic vulnerability. Full-time workers (32.6%) had slightly lower positivity, possibly due to more stable employment with structured safety protocols or remote work options. Homemakers (19.4%) and students (17.1%) had the lowest infection rates, consistent with reduced occupational exposure and greater ability to quarantine (Fig 3C). These trends indicate that unstable employment, lack of paid sick leave, and absence of workplace protections may heighten exposure among Latino populations employed in essential sectors such as agriculture, food service, construction, and custodial labor.

#### 3.4. Vaccine Concerns and Community Trust

Latino participant vaccine attitudes reflected safety concerns, misinformation, and institutional mistrust (*Fig 3D*). Fear of side effects or doubts about vaccine safety, reported by 1,580 participants, highlighting enduring influence of misinformation and limited access to culturally relevant health communication. Media influence was substantial; 1,100 participants indicated that their views were shaped by media coverage, often inconsistent or sensationalized early in the pandemic. Similarly, 1,050 participants were unsure of the vaccine’s effectiveness, and 980 participants believed it had been developed too quickly, reflecting both scientific uncertainty and communication gaps around vaccine trials and approval processes.

Trust and social concerns were also substantial (*Fig 3D*). Participants cited distrust of the healthcare system or government (580 participants), reflecting longstanding inequities in healthcare access and treatment that shape vaccine hesitancy in Latino communities. A smaller subset of participants raised religious (350) or personal beliefs (200 personal freedom; 200 vaccine not needed) as factors influencing their decision, while 250 participants said they did not know what the vaccine contained, suggesting that knowledge gaps could be addressed through accessible education campaigns. Overall, these findings demonstrate that vaccine hesitancy among Latino populations is multifactorial, influenced by safety and efficacy concerns and institutional distrust.

### 4. Biomarker Discovery

#### 4.1. Putative biomarker extraction by secondary use of peripheral blood RNA-seq data from pediatric MIS-C patients

We utilized RNA-seq data from 217 whole blood specimens collected under the RADx-rad program from SARS-CoV-2 patients presenting with varying MIS-C severity (7). This secondary analysis had two complementary objectives: i) to identify sets of highly discriminant expressed genes that characterize specimens among latent groupings of transcriptional similarity, and ii) to assess whether those groups aligned with reported MIS-C severity (*Fig 4A*).

**Figure 4.**
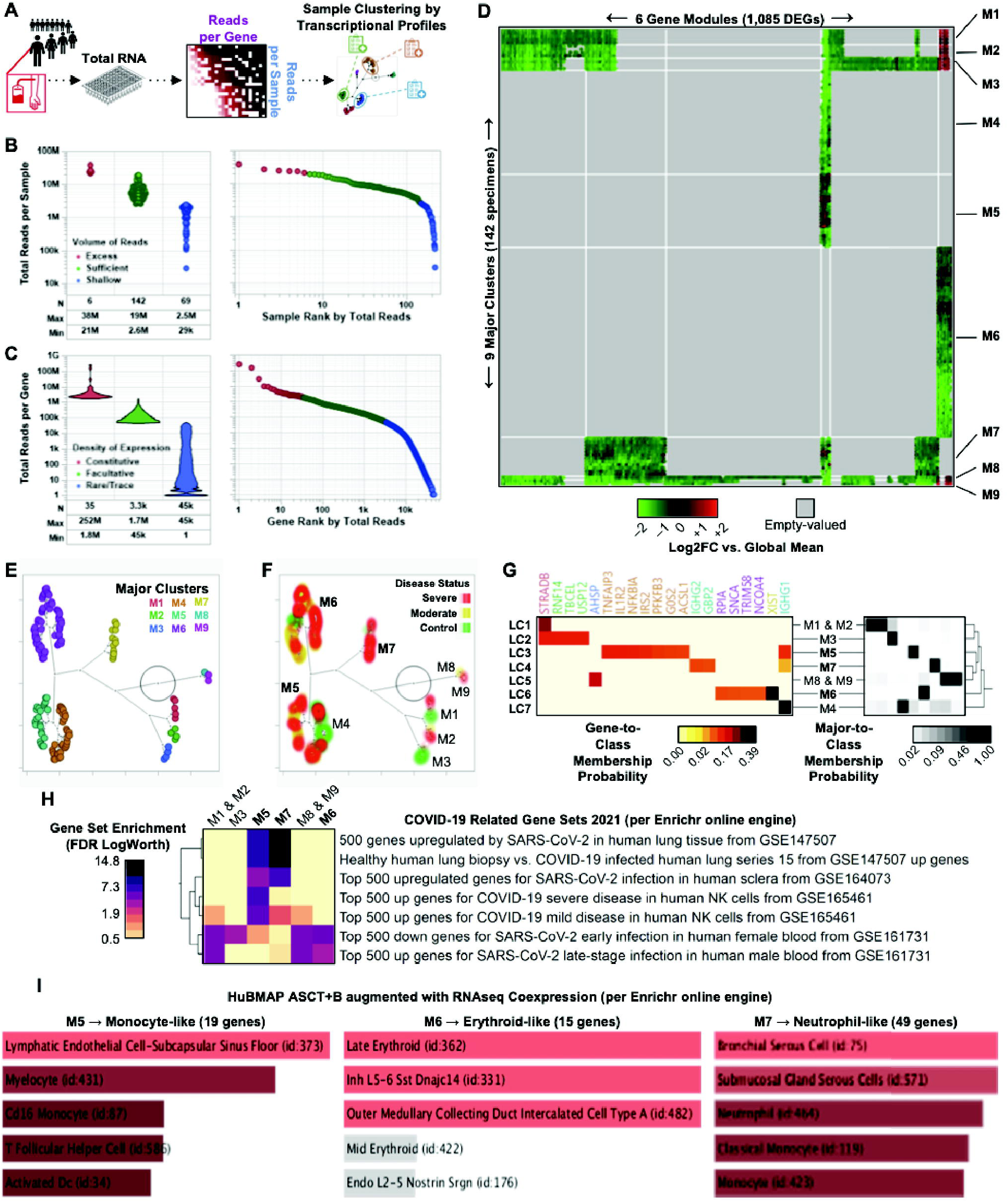
Identification of biomarkers of COVID-19/MIS-C severity in children by secondary analysis of sparse RNA-seq data from whole blood specimens. A. Schematic of RNA-seq analytical strategy to coalesce unsupervised clustering of participant transcriptomes with clinical metadata from participants in phs002781 study. B. Identification of subsample of 142 specimens to include in secondary analysis based on comparability in volume of reads sufficiency. C. Identification of 3,349 facultative genes of interest to leverage for expression pattern recognition by unsupervised clustering based on density of expression. D. Specimen × Gene heatmap of gene expression fold differences with respect to the global mean of all specimens, ordered by patterns of expression similarity grouped into gene modules (x-axis) and major sample clusters (y-axis: M1 through M9). E. Plot of 2D projection of the hierarchical clustering dendrogram of specimens, with coloring to indicate major cluster assignments. F. Overlay of non-parametric density contours of disease severity onto constellation plot of specimens, with inset labeling to indicate major cluster assignments. G. Hierarchical clustering of estimated item-response probabilities via latent class analyses for a subset of 20 top-scoring candidate biomarkers (left, heatmap: yellow to red gradient) and all 9 major specimen clusters (right, heatmap: white to black gradient) that correspond with 7 latent classes. H. Heatmap of enrichment significance for gene sets in the “COVID-19 Related Gene Sets 2021” database from Enrichr using 100 top-scoring candidate biomarkers corresponding with major specimen clusters per latent class analysis. I. Bar graphs of top 5 enriched gene sets in the “HuBMAP ASCT+B augmented with RNAseq Coexpression” database from Enrichr using subsets of genes from 100 top-scoring biomarkers which correspond with major clusters of specimens M5-M7 and overlap with study participants presenting severe COVID-19/MIS-C.

Specimen libraries showed variable effective transcriptome representation, producing a sparse, under-sampled cross-study gene expression matrix (see *Materials and Methods*). The tallies of aligned read counts per gene were also heavily unbalanced across individual specimens, despite deriving from standardized alignment and bioinformatics workflows (7). We reasoned that the undersampled expression matrix in our study was similar ultra-sparse scRNA-seq cell×gene datasets, so we could identify specimens with poor transcriptome readout efficiency to discard from our analysis (8). This strategy yielded 6 specimens with excess data volume, 142 with sufficient data volume, and 69 with shallow data volume (*Fig 4B*) (see *Materials and Methods*).

Based on gene expression matrix distribution features reminiscent of single-cell RNA-seq data footprints, we adopted the SALSA analytical workflow (see *Materials and Methods*) (8), which treats specimen-level data similarly to single-cell approaches, and was previously established to retrieve candidate biomarkers from sparse and untargeted RNA-seq data (8,10). This also allowed us to reduce the analytical search space for unsupervised biomarker extraction by focusing statistical analysis on agnostically determined facultative genes. We focused our unsupervised clustering analysis to ∼3,300 facultative genes (*Fig 4C*) and applied dimensional reduction of gene expression patterns via unsupervised clustering. In all, we identified 9 major specimen clusters (M1 through M9), driven by expression patterns across 1,085 differentially expressed genes (DEGs) grouped into 6 distinct gene expression modules. Sorting data by cluster and gene-module memberships confirmed that the 142 sufficient specimens formed a sparse specimen×gene expression matrix (*Fig 4D*).

#### 4.2. Clustering of specimens by severity

We examined whether major specimen clusters overlapped with distinct MIS-C severity patterns. We mapped reported MIS-C severity levels onto their corresponding specimens and observed that severe MIS-C mapped heterogeneously, but concentrated within a subset of major clusters, especially specimens from M5, M6, and M7 (*Fig 4E*). Among the 1,085 DEGs, we identified candidate expression biomarkers most closely associated with major clusters. This analysis yielded a minimal assignment probability baseline of 6.37×10^-5^, with 135 DEGs having assignment probabilities greater than the baseline to at least one cluster. We excluded genes with a maximum assignment probability across clusters less than a random assignment threshold of 1 in 270, resulting in 100 top-scoring candidate biomarkers. These biomarkers tracked almost exclusively with one of seven latent gene expression classes (LC1 through LC7), each corresponding to one or two combined major specimen clusters. We used two-way unsupervised hierarchical assignment score clustering, which helped identify a smaller subset of 20 top-scoring biomarkers with the sharpest latent class membership probabilities by visual inspection (*Fig 4G*).

Specimens from patients with severe MIS-C predominantly mapped to major clusters M5 through M7 (*Fig 4F*). Established correspondence between major clusters and latent gene expression classes (M5 → LC3, M6 → LC6, and M7 → LC4, as shown in *Fig 4G*), linked specimen donors with an enrichment of expression for distinct biomarker genes characteristic of specimen groupings. We performed gene set enrichment analysis on the 100 candidate biomarkers, grouped by their distinct latent class associations, to assess i) whether biomarkers linked to specimens from donors with severe MIS-C reflected previously reported gene expression findings in SARS-CoV-2-infected human primary cells and tissues; and ii) whether severe MIS-C-associated gene sets represented enriched responses from specific anatomical structures, cell types, or biomarkers (see *Materials and Methods*).

#### 4.3. Cluster Characterization

Using Enrichr (11) we found that specimens exhibited varying informative biomarker potential levels (Table 2). The M5, M6, and M7 clusters had transcriptional indicators of immune/blood, optical, or lung systems (Fig 4H-I).

**Table 2.**
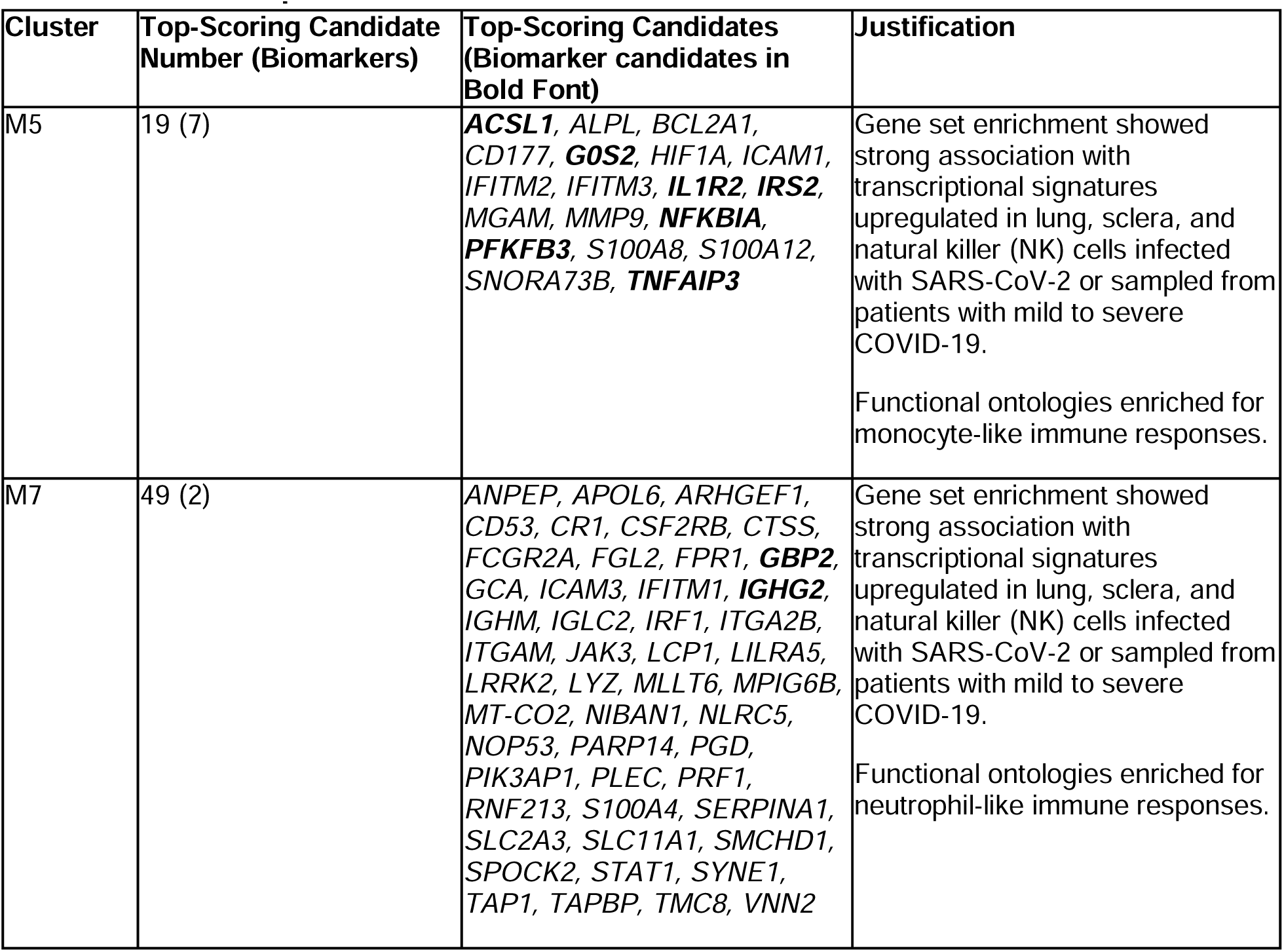
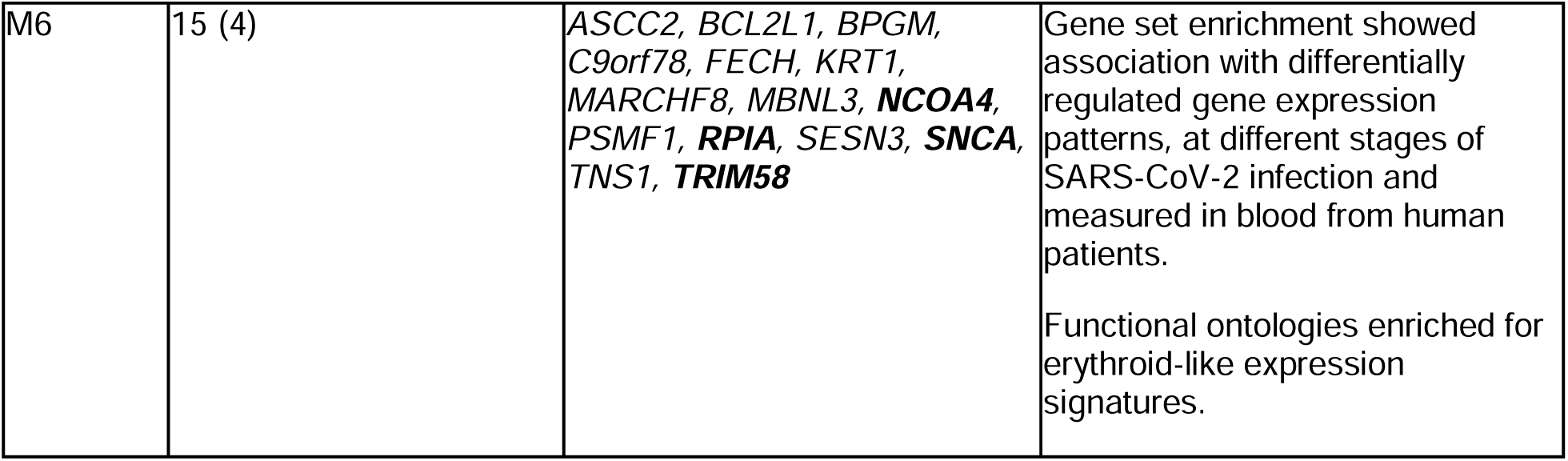
Biomarker potential of Candidates.

Our secondary analysis shows that blood specimens from patients with severe MIS-C display immune signatures at the gene expression level consistent with enhanced phagocytic activity (as in M5 and M7 genes), suggesting active hemolytic reactions in a subset of patients (as in M6 specimens). These features can be identified through first-tier differential expression screening using at least three of thirteen measurable biomarker genes.

### 5. Structural Hardship, Educational Attainment, and COVID-19 Vaccination: A Confirmatory Factor Analysis of Precarity

#### 5.1. Study of Structural Barriers in Vaccine Uptake

We analyzed pooled data from nine RADx-UP studies accessed through the Data Hub to evaluate how cumulative household hardship interacts with educational attainment to shape COVID-19 vaccination uptake. Together, these projects contributed data from 5,397 adult participants across diverse populations and settings, including urban underserved communities, syringe-exchange clients, medically underserved adults, and caregivers of children with intellectual and developmental disabilities. The Data Hub’s standardized architecture enabled consistent vaccination status, demographic, and hardship indicator measurement across studies. The combined sample was demographically diverse (53% female, 47% male, with a mean age of 49 years), with 45% reporting education beyond high school. Overall, 76% of participants were vaccinated against COVID-19, though coverage varied across populations.

#### 5.2. Household Precarity Factor

To quantify cross-study cumulative household hardship, we applied a Confirmatory Factor Analysis (CFA) on six harmonized hardship indicators available through the Data Hub’s Common Data Elements (12–14). These indicators included limited healthcare access, unstable housing, food insecurity, lack of clean water, difficulty obtaining medications, and limited transportation. All six ordinal variables were measured consistently across studies due to Data Hub harmonization, allowing them to be modeled jointly as observed indicators of a shared latent construct. As shown in Fig 5A, most participants reported no challenges, though minor and major challenges were common, particularly for food security, transportation, and access to healthcare and medications. These distributions provided the empirical basis for modeling cumulative hardship using a latent factor approach.

**Figure 5.**
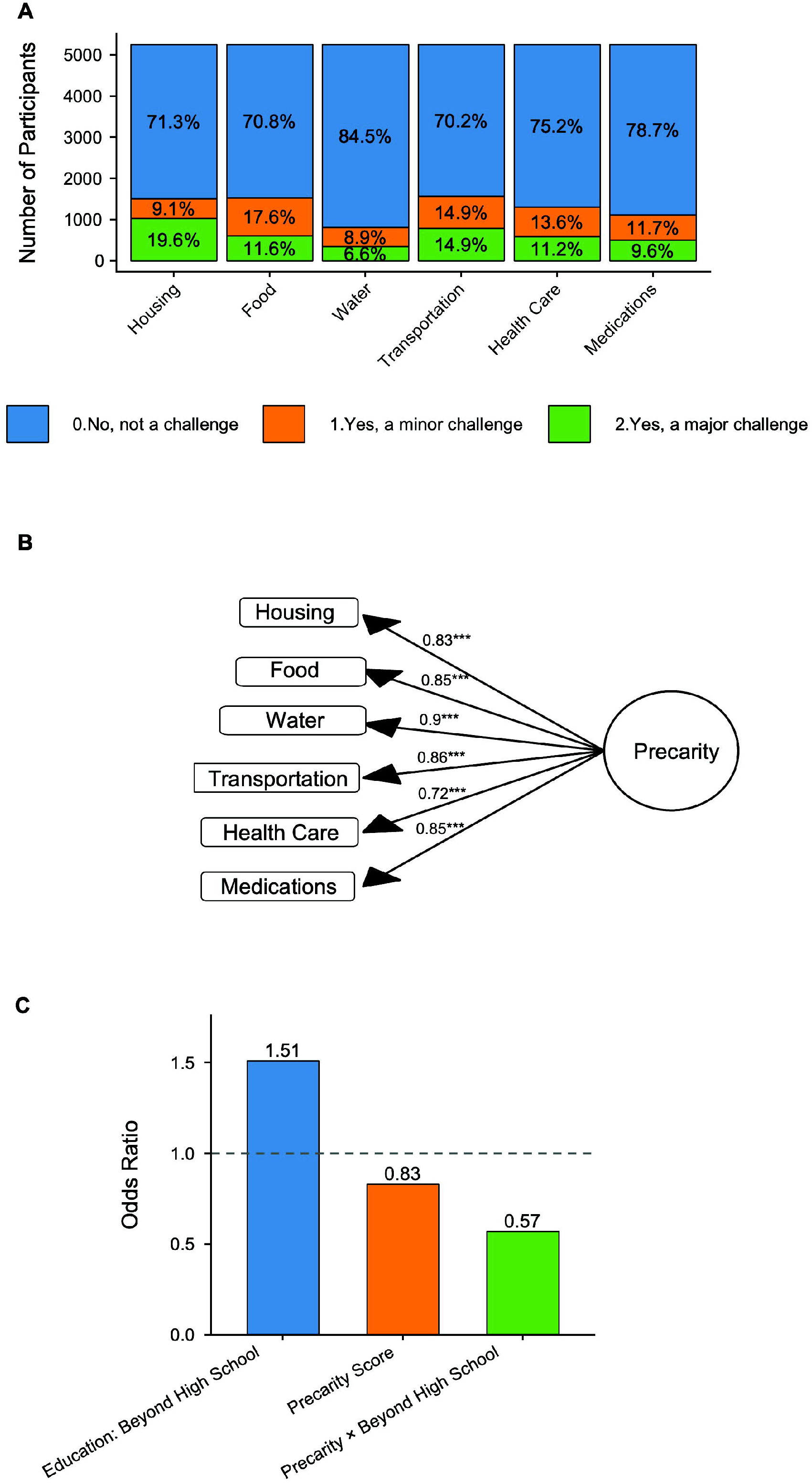
The Paradox of Education: How Structural Barriers Shape Vaccine Uptake. Panels A-C show how structural hardship interacts with education to influence COVID-19 vaccination uptake. A. The prevalence of six household challenges, healthcare, housing, food, water, medication, and transportation, that together form the foundation of the Precarity construct. B. The Confirmatory Factor Analysis (CFA) model combines these indicators into a single Precarity score (factor loadings = 0.72-0.90). C. Adjusted odds ratios demonstrating that individuals with greater household precarity were less likely to be vaccinated, and that high precarity substantially reduced the protective effect of higher education. Data are drawn from 5,397 participants across nine RADx-UP studies representing diverse underserved U.S. populations.

The CFA results showed that all six indicators loaded strongly on a single latent household Precarity factor (standardized loadings λ = 0.72 to 0.90). Model fit was excellent (χ²[9] = 48.2, *p* < 0.001; CFI = 0.998; TLI = 0.996; RMSEA = 0.046). Higher factor scores reflected greater cumulative hardship and varied substantially across studies, geographic regions, and educational levels. This validated Precarity score provided a unified structural hardship measure across the RADx-UP projects and served as the primary independent variable for evaluating its relationship with COVID-19 vaccination uptake in downstream regression models (Fig 5B).

#### 5.3. Vaccination Uptake Model

We estimated a multivariable logistic regression model to evaluate how household precarity and educational attainment jointly predict COVID-19 vaccination uptake, adjusting for age, age-squared, sex at birth, substance use risk, and health insurance status. Educational attainment was represented using two indicator variables: “high school/GED” and “beyond high school”, with “less than high school” as the reference category.”

Greater household precarity was associated with lower vaccination odds (OR = 0.83, *p* < 0.10), indicating reduced uptake as structural hardship increased. Participants with education beyond high school had substantially higher odds of vaccination (OR = 1.51, *p* < 0.001), while those with only a high school/GED resembled the reference group (OR = 0.97). The interaction between precarity and education revealed a key moderating effect. The Precarity × Beyond High School term was statistically significant (OR = 0.56, *p* < 0.001), demonstrating that the protective effect of higher education diminishes sharply as household hardship increases (Fig 5C).

## Discussion

The Data Hub was designed to be an exemplar of an integrated, FAIR-compliant data platform, demonstrating how centralized and harmonized repositories can accelerate research for future public health challenges. We independently investigated diverse uses, and found that the Data Hub, a central repository with an integrated analysis platform (2) seamlessly facilitated secondary research. The program’s harmonized datasets provide a foundation not only for the current pandemic response but also for future-ready diagnostic and surveillance systems.

First, using hub-based RADx-rad data, we found that cutting-edge diagnostic methods can be developed and validated across diverse specimen types and viral variants. The Data Hub’s harmonized datasets enabled systematic, side-by-side innovative technology comparisons, demonstrating that RADx-rad transformed infectious disease diagnostics, enabling rapid, accurate, and scalable detection. Moreover, the Data Hub’s secure workspace provided integrated tools for data querying, analysis, and machine learning, which facilitated reproducibility and accelerated discovery.

Secondly, we found that wastewater-based epidemiology (WBE) provided early, unbiased detection of SARS-CoV-2 trends, often anticipating clinical activity, particularly around semester transitions (4). Comparing NC A&T’s dorm-level campus data with a Miami University’s Data Hub-based community-scale showed that catchment scale differences (dorm vs. community) explained amplitude/variance differences and underscored the complementarity of local and regional surveillance layers: local data are better for action; regional data are better for context.

Consistent with clinical and epidemiologic studies, we confirmed that the Delta variant was associated with higher clinical severity, while Omicron lineages (BA.1/BA.5/XBB) exhibited greater transmissibility and immune escape, driving rapid campus surges despite lower per-infection severity than Delta (5,6). Operationally, because faster growth and immune escape can stress campus operations (absenteeism, isolation housing) even when severity is lower, WBE can help to detect and compare these dynamics in near-real time for targeted mitigation (4).

We noted that interpretation is affected by low-copy samples near the LoD, occasional temporal gaps, evolving clinical testing volumes, plumbing/site differences, and incomplete flow/population metadata. To improve cross-site comparability in future work, we recommend integrating normalization using pepper mild mottle virus (PMMoV) (15), expanding surveillance to RSV/Influenza/AMR targets, and adopting dashboards that link wastewater, clinical, and mobility data for campus-level decision support (9). Importantly, this analysis also shows that Data Hub wastewater studies can serve as a reference layer when a campus already has local wastewater but wants to understand whether its patterns match a broader community signal.

Regarding health disparities, we found evidence supporting prior observations that the COVID-19 pandemic disproportionately affected marginalized United States communities, with Latino populations experiencing some of the highest rates of infection, hospitalization, and mortality.

By integrating findings from six RADx-UP community intervention datasets across multiple U.S. states to examine how social determinants and community engagement strategies shaped COVID-19 outcomes among Latino populations, we found that that lower educational attainment, unstable employment, and lack of insurance were strongly associated with higher infection rates. These findings support the hypothesis that structural and social determinants of health contribute substantially to Latino community infection risks and show the enduring influence of structural inequities on health.

Vaccine hesitancy was a persistent barrier, driven primarily by safety concerns, perceived vaccine development speed, and government institution distrust. Prior work in 2020 indicated that Latino communities experienced a 54% excess mortality compared with 12% among non-Hispanic Whites, and infection rates were over three times higher than the general population (16). Vaccination rates lagged behind, with only 32% of Latinos vaccinated by May 2021 compared with 43% of non-Hispanic Whites (17). These disparities were partially influenced by social determinants of health, including limited healthcare access, employment instability, educational inequities, and structural barriers that reduce prevention and care opportunities. Strengthening vaccine confidence requires transparent communication, community-driven outreach, and linguistically and culturally tailored messaging delivered by trusted local voices (18). These results reinforce the need for health equity frameworks that address upstream determinants such as employment, education, and healthcare access, while leveraging community trust to deliver public health interventions.

Nevertheless, this health disparity analysis is descriptive and cross-sectional, limiting the ability to infer causality or temporal relationships between socioeconomic factors and infection risk. Self-reported data may introduce recall or reporting bias, particularly regarding COVID-19 testing and vaccination attitudes. The sample may also not represent all Latino subpopulations, and state-level differences were not statistically adjusted. Despite these limitations, the findings provide valuable insight into disparity patterns that warrant further investigation. Future research should include longitudinal monitoring and multisite collaborations to assess community partnership durability, social media-based outreach impacts, and promotoras-led intervention scalability for other minority populations. Mixed-method approaches integrating behavioral, social, and contextual data will be critical for developing sustainable, inclusive strategies that strengthen community resilience beyond the COVID-19 pandemic and ensure more equitable responses to future public health emergencies.

SARS-CoV-2 infections can elicit both COVID-19 in adults and MIS-C in children, but these two complex diseases are often researched and managed separately, impeding effective disease detection and monitoring. This has slowed the discovery of novel actionable biomarkers and phenotypic traits applicable to both diseases. As such, our understanding of commonalities between COVID and MIS-C molecular physiology remains limited. Using Data Hub-based multi-modal research data from adults and children diagnosed with COVID and/or MIS-C, we evaluated potential indicators of SARS-CoV-2 infection that may predict COVID-19 or MIS-C severity starting from clinical or molecular patient data.

The PreVAIL kIds cohort, a translational research consortium of eight independently-run NICHD-led clinical studies with roughly 10,000 pediatric participants (19), offers molecular profiling data amenable to multi-modal integration and biomarker discovery. Using whole blood RNA-seq data from one of these studies (7), we found strong associations between MIS-C severity and highly enriched immune-related transcriptional profiles, linking clinical observations to underlying physiological mechanisms. These results posit testable molecular biomarker candidates to forecast MIS-C incidence, severity, and progression across the SARS-CoV-2 infection and host response timeline. By amalgamating physiological and clinical traits across severity levels, our secondary analysis reveals new analytical strategies to project multisystemic disease severity in patients presenting with early symptoms or clinical SARS-CoV-2 infection diagnoses.

The biggest challenge in processing these datasets came from disparities between total sequenced and aligned read counts, possibly due to differentially saturated sequencing libraries. To address this, we adopted a statistical workflow tailored for sparse expression datasets (i.e., single-cell RNA-seq analysis) that pre-emptively mitigated statistical bias, prioritized robustly measured gene expression levels, and optimized latent association discovery between observed clinical phenotypes and molecular data profiles in a streamlined fashion.

Finally, we show how the centralization of data within the Data Hub can enable equity-focused research by harmonizing complex, multi-site data for advanced analytic modeling. The Data Hub’s harmonized datasets, transparent metadata, and analytic infrastructure enabled reproducible cross-cohort modeling. Using CFA, we derived a unified Precarity score that quantifies cumulative household hardship, a key dimension of structural vulnerability affecting COVID-19 vaccine uptake.

The results reveal a central paradox: while education is typically protective for health behaviors, its benefit diminishes when structural barriers are high. Individuals with higher education but greater precarity were substantially less likely to be vaccinated. This pattern supports a polysocial risk framework, which recognizes that overlapping social risks including housing, transportation, and healthcare access, operate collectively rather than in isolation. From a policy perspective, these findings suggest that interventions addressing upstream structural hardships, rather than focusing solely on education or awareness, may yield greater, more equitable gains in vaccine coverage.

While the Data Hub was effective in harmonizing and centralizing RADx Data, investigators should be aware of some challenges. These include data missingness, including incomplete or inconsistent demographic/clinical information and residual variability in study design and patient populations. While adopting metadata and FAIR-aligned ontologies partially alleviates these issues, further standardization may be required based on investigators’ study questions. Data access and governance frameworks can also limit rapid data access and cross-institutional analyses. Implementing interoperable governance models and harmonization policies across NIH-supported repositories could reduce such bottlenecks. Moreover, future work linking RADx data (acute COVID infection data) with initiatives such as REsearching COVID to Enhance Recovery (RECOVER; long COVID) (20), may facilitate investigations into long-term sequelae of SARS-CoV-2 infection, while integrations with All of Us (21) or NHLBI’s BioData Catalyst (22) could accelerate broader research on health disparities, biomarker trajectories, and social determinants of health on infectious disease outcomes.

## Data Availability

Data used in this study are available from the RADx Data Hub.

https://radxdatahub.nih.gov/

## Acknowledgements

We thank the participants of RADx-funded studies, the RADx Data Hub data generators and coordinators including RADx study investigators, and the (C)DCCs that provided the foundation that enabled this work. We thank Marcos Martinez-Romero, Alissa Fujimoto, Nilesh Mistry, Ashley Sier, Aubrie Weyhmiller, Aria Henry, and Ashley Evans, and Leah Mason, for technical assistance and operational support in the execution of this work. We thank current and former RADx Data Hub Partners for their efforts on the RADx Data Hub.

## Author Contributions

P.W.R., T.R., N.M., O.A.L., L.J.F., and K.N.S. wrote and revised the manuscript. M.D.K., M.U.A., J.S., and B.F. provided critical content input and feedback, and reviewed the manuscript. M.A.K., M.A.M., S.S., and A.K. provided support as principal investigators of the RADx Data Hub.

